# Characteristics of effective interventions to optimise retention for early career nurses: a scoping review

**DOI:** 10.1101/2024.12.18.24319204

**Authors:** Krishangi White, Adrian Goldsworthy, Sarah Bateup, Neil Meigh

## Abstract

**Background:** Nursing shortages, driven by high turnover rates among early career registered nurses (ECNs), present a critical global challenge, threatening workforce sustainability, compromising patient care quality, and imposing financial burdens on healthcare systems. While various interventions have been developed to improve retention, gaps in reporting, scalability, and long-term evaluation hinder their widespread adoption. This review applies Proctor et al.’s implementation science framework to evaluate the design and operationalisation of retention strategies, addressing existing gaps and identifying actionable insights.

**Objective:** This scoping review synthesises evidence on the characteristics of effective interventions designed to enhance retention and reduce turnover among ECNs.

**Inclusion criteria:** This review included studies evaluating interventions aimed at improving retention or reducing turnover of ECNs within the first five years of practice. Eligible studies presented original data or systematic reviews addressing intervention characteristics and outcomes.

**Methods:** The review adhered to the JBI methodology for scoping reviews and PRISMA-ScR guidelines. Systematic searches of Medline, EMBASE, PsycINFO, CINAHL, and the Cochrane Library were conducted in May 2024. Articles published in English were screened independently by two reviewers, with data extraction and synthesis guided by Proctor et al.’s implementation science framework. Results were synthesised narratively and presented in tabular formats.

**Results:** From 3,462 records, 21 studies met inclusion criteria. Interventions varied widely, including mentorship (n = 9), preceptorship (n = 8), in-person classes (n = 10), clinical simulations (n = 6), reflective practice, and career advancement programs. Program durations ranged from 8 weeks to 2 years, with one year being the most common. Factors associated with success included tailored content for ECNs, structured competency frameworks, mentor training, and integration into organisational leadership and culture. Challenges such as variable reporting standards and inconsistent evaluation methods were noted.

**Conclusions:** Effective retention strategies for ECNs require structured, supportive, and competency-based interventions tailored to organisational and individual needs. Emphasising mentor preparation, career development, and specialised approaches for high-stress environments, such as critical care, can enhance program outcomes. Improved reporting standards and methodological rigour are necessary to scale and adapt these programs across healthcare systems, ultimately contributing to a more stable and resilient nursing workforce.

## Introduction

Nursing shortages represent a critical global challenge, exacerbated by persistently high turnover rates among newly graduated and early-career nurses (ECNs). These shortages exacerbate staffing challenges, compromise the quality of patient care, and incur substantial financial costs for healthcare organisations. The average cost of turnover for a bedside registered nurse in the United States is estimated at USD $52,350, with hospitals experiencing annual losses between $6.6 million and $10.5 million due to inefficiencies related to turnover (1,2). In Australia, nurses and midwives account for 54% of the health workforce, yet projections indicate a shortfall of 123,000 nurses by 2030, underscoring the unsustainable nature of current trends (3). Alarmingly, a recent Queensland survey revealed that 46% of nurses and midwives are considering leaving the profession within the next 12 months, highlighting the urgency of addressing retention challenges (4).

The first five years of a nurse’s career are widely recognised as a pivotal period, characterised by disproportionately high turnover rates due to challenges such as job dissatisfaction, burnout, and limited professional support (1,5,6). Early-career nurses frequently experience ‘transition shock,’ a phenomenon characterised by challenges such as adapting to professional practice, high workloads, and inadequate mentorship (7,8). The COVID-19 pandemic has further exacerbated these stressors, introducing moral injury and heightened burnout among ECNs as they navigate increasingly demanding and resource-constrained environments (9). The complexity of care delivery in high-acuity environments, combined with organisational stressors such as inadequate staffing, poor leadership, and unhealthy workplace cultures, further intensifies attrition risks (10–12).

Definitions of “early-career” or “new graduate” nurses vary significantly in the literature. For instance, Brown et al. (13) define early-career nurses as those in their first three years of practice, while Vázquez-Calatayud et al. (6) focus on newly graduated nurses within their first 12 months. Such variability complicates the design and evaluation of interventions aimed at retention, as different studies address distinct phases of early professional development. This review adopts a broader definition, encompassing the first five years of practice to account for the extended period during which transition challenges persist. This approach aligns with evidence suggesting that the risk of attrition remains high throughout these early years, particularly in hospital settings (5,14).

Healthcare organisations have implemented various interventions to address these challenges, including structured mentorship programs, residency models, clinical simulations, and reflective practices. Transition-to-practice programs, such as the UHC/AACN model, focus on bridging the gap between education and practice by providing tailored mentorship, competency-based learning, and structured feedback mechanisms (8,15). Mentorship and preceptorship initiatives, such as the Norton Navigators program, demonstrate the critical role of supportive relationships, organisational leadership, and structured feedback in fostering ECN retention (11,16). Evidence highlights that interventions tailored to the specific demands of high-stress settings, such as critical care or rural hospitals, can significantly enhance retention by addressing the unique challenges faced by nurses in these contexts (10,17). Wellness initiatives and flexible scheduling, particularly in response to the COVID-19 pandemic, further underscore the importance of adaptable and supportive work environments (9).

Despite these advances, gaps persist in the standardisation and reporting of intervention characteristics. Existing literature frequently lacks consistency in defining intervention components, such as their theoretical underpinnings, implementation processes, and long-term impacts (5,14). Emerging issues, such as the integration of AI and technology, offer opportunities to enhance workflows and job satisfaction. However, these developments also raise concerns about the need for ongoing training and adaptation, which can further burden ECNs if not managed thoughtfully (9,12,18).

To address these gaps, this scoping review applies Proctor et al.’s (19) implementation science framework, which provides a systematic approach to evaluating intervention characteristics. This framework identifies critical dimensions such as “actors” (who implements the intervention), “actions” (what is done), “targets” (who is affected), “dose” (intensity and frequency), and theoretical justification. By mapping these elements, this review aims to provide actionable insights into the design and execution of effective retention interventions for ECNs. The adoption of this framework ensures that the findings are not only theoretically grounded but also practical and replicable across diverse healthcare settings. No current or in-process scoping reviews that captured Proctor’s implementation characteristics were identified.

Retention of ECNs is essential for ensuring workforce sustainability, improving patient outcomes, and reducing the financial burden on healthcare organisations. By synthesising evidence using an implementation science lens, this review aims to provide a replicable blueprint for designing and scaling effective retention strategies. This work contributes to addressing the nursing workforce crisis and fostering a resilient and sustainable healthcare system.

## Methodology

### Protocol and Registration

This scoping review was conducted in accordance with the JBI methodology for scoping reviews (20) and is reported in accordance with the Preferred Reporting Items for Systematic Reviews and Meta-Analyses extension for scoping reviews (21). The protocol was pre-registered on the Open Science Framework (https://osf.io/k389h) on 07/05/2024.

### Eligibility Criteria

A pre-determined eligibility criteria was developed informed by the population (registered nurses within the first 5 years of registration), concept (retention strategies) and context (hospital settings). A narrow definition of registered nurse (RN) was utilised encompassing those individuals entering the workforce for the first time with an undergraduate nursing degree. Advanced practice nurses such as nurse practitioners and clinical nurse specialists in addition to assistants in nursing (AIN) enrolled nurses (EN) and endorsed enrolled nurses (ENN) were excluded due to the role’s distinct differences in scope of practice and responsibility from registered nurses. In recognition of the unique challenges new graduate and early career nurses face, as well as the high rate of attrition this population displays RNs with greater than five years of experience as well as student nurses and interns were similarly excluded. Additionally, articles which did not evaluate or report improvements in RN retention or failed to improve retention were excluded. Due to resource constraints, only articles published in English were included.

### Search Strategy

A search strategy was developed utilising a three-step methodological approach with the assistance of a health sciences research librarian (SB). Initially, a preliminary search was developed to identify seed articles within PubMed. Secondly, results were reviewed to a identify additional search terms. The final search strategy was translated for additional databases using the Systematic Review Accelerator Polyglot Search tool (22) with subject headings being updated by hand (Appendix 1).

### Information Sources

Seven databases (PubMed, Embase, CINAHL, PsycINFO, Scopus, Web of Science) were searched on May 10th, 2024. No date or language limits were applied. Results from database searches were exported into Endnote X9 (23).

### Selection of Sources of Evidence

Duplicate results were removed using Covidence’s automated deduplication software (24). Screening was subsequently undertaken within Covidence by two authors (KW, AG) first by title and abstract and then by full text with discrepancies resolved through unanimous agreement.

### Data Charting Process

A draft extraction table was developed within Microsoft Excel (25) prior to data extraction to align with the questions of the scoping review informed by the implementation science framework described by Proctor et al. (19). Final extraction of data was undertaken and manually compiled by two authors (KW & AG).

### Synthesis of Results

Data was synthesised and has been displayed in both figures and tables, accompanied by a written description. Data pertaining to section one (name it) and three (specify it) of the framework described by Proctor et al. (19), including the actor, action, action target, temporality and dose and implementation was extracted and tabulated. Within this framework the actor refers to the individual(s) who enacts the strategy, the action refers to the steps undertaken. The action target was extracted and synthesised to capture data relating to specific cohorts of new graduate and early career RN entering within specific setting within the hospital. The temporality refers to the overall length of time the program was delivered whilst the dose refers more specifically to the frequency and length on individual intervention components (e.g. support sessions, workshops). Additionally, the empirical, theoretical or pragmatic justification for the choice of interventions and implementation strategies was tabulated. To assist readers with reoccurring descriptions, definitions of interventions are provided in Table 1.

**Table 1:** Definitions of common interventions.

| Intervention Name | Definition |
| --- | --- |
| Preceptor | An experienced nurse providing direct supervision, guidance, and support to newly graduated nurses during their clinical transition (26). |
| Mentor | An experienced nurse offering long-term career guidance, emotional support, and professional development to less experienced nurses (27). |
| Clinical coordinator | A healthcare professional responsible for coordinating clinical activities, ensuring efficient operation of clinical services, and supporting both staff and patients to maintain high standards of care (28). |
| Clinical nurse educator | A registered nurse responsible for designing, implementing, and evaluating educational programs to ensure high clinical practice standards (29). |
| Volunteer nurse ambassador | A retired nurse returning to support and mentor newly graduated nurses, enhancing their communication skills and confidence (30). |
| Nurse navigator | A nurse specialising in guiding patients through the healthcare system and supporting new graduates in their professional integration (8). |
| Program coordinator | Oversees nurse residency programs, ensuring curriculum development, training coordination, and support for preceptors and new graduates (31). |

## Results

### Selection of Sources of Evidence

Database searching led to the retrieval of 3,462 articles of which 1,732 were automatically removed as duplicates. Title and abstract screening of the remaining 1,730 articles led to the exclusion of 1,652 articles. Full-text screening of the remaining 78 articles was undertaken with substantial agreement between authors (Cohen’s Kappaϑ= .744) (24). Following the exclusion of a final 57 articles, 21 articles were included within the scoping review (Figure 1).

**Figure 1:**
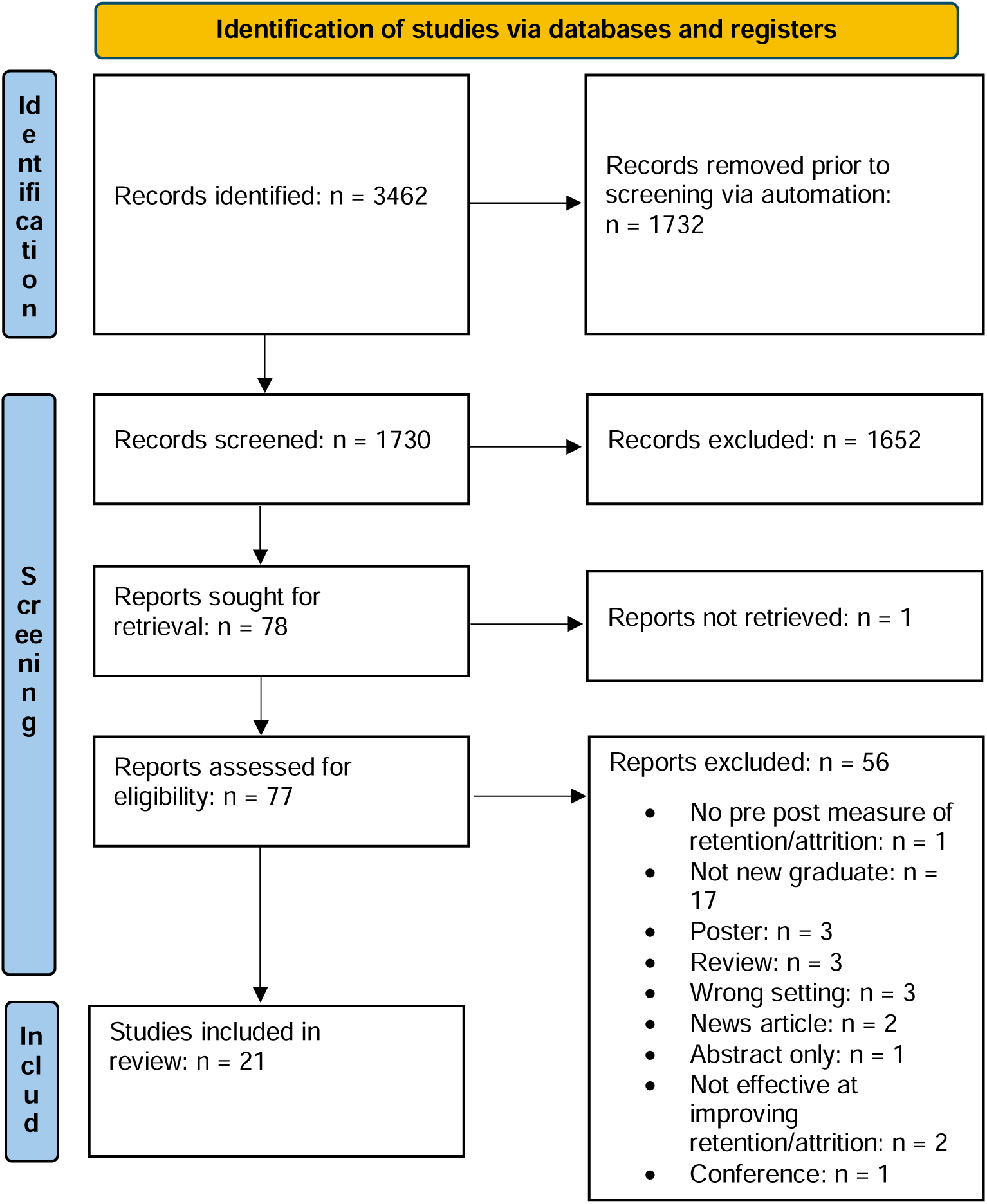
PRISMA flow diagram

### Overview

The 21 included studies were undertaken in three countries; the United States of America [(8,15,18,26–41)] Canada [(42)] and the United Kingdom [(43)] (Figure 2). Notably only two articles were published since the outbreak of COVID-19 [(39,40)]. Interventions to improve NGN retention were found to be delivered primarily within tertiary facilities with only one article representing a primary healthcare setting. A variety of tertiary facilities and departments were represented including children’s hospitals [(36,38)], critical care hospitals [(34,40,42)], rural community hospitals and community hospitals [(28)]. Whilst some interventions focused on more focused interventions such as touchpoints [(39)], many interventions consisted of a broader nursing retention strategy commonly, referred to as residencies [(15,31,33,35,42)] or fellowships [(34)], that consisted of multiple elements. For example, many of these broader strategies included a combination of orientation, mentorship and/or preceptorship (Table 2).

**Figure 2.**
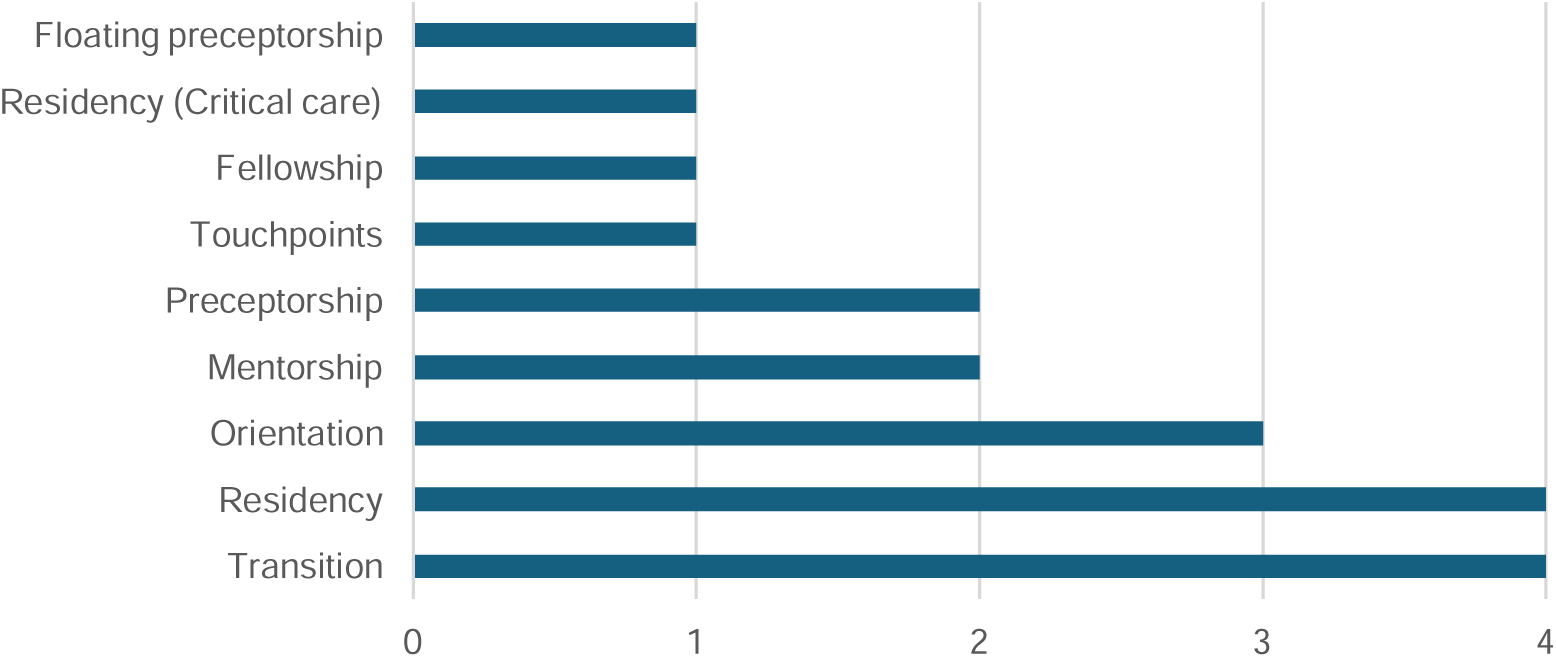
Names of interventions

**Table 2:**
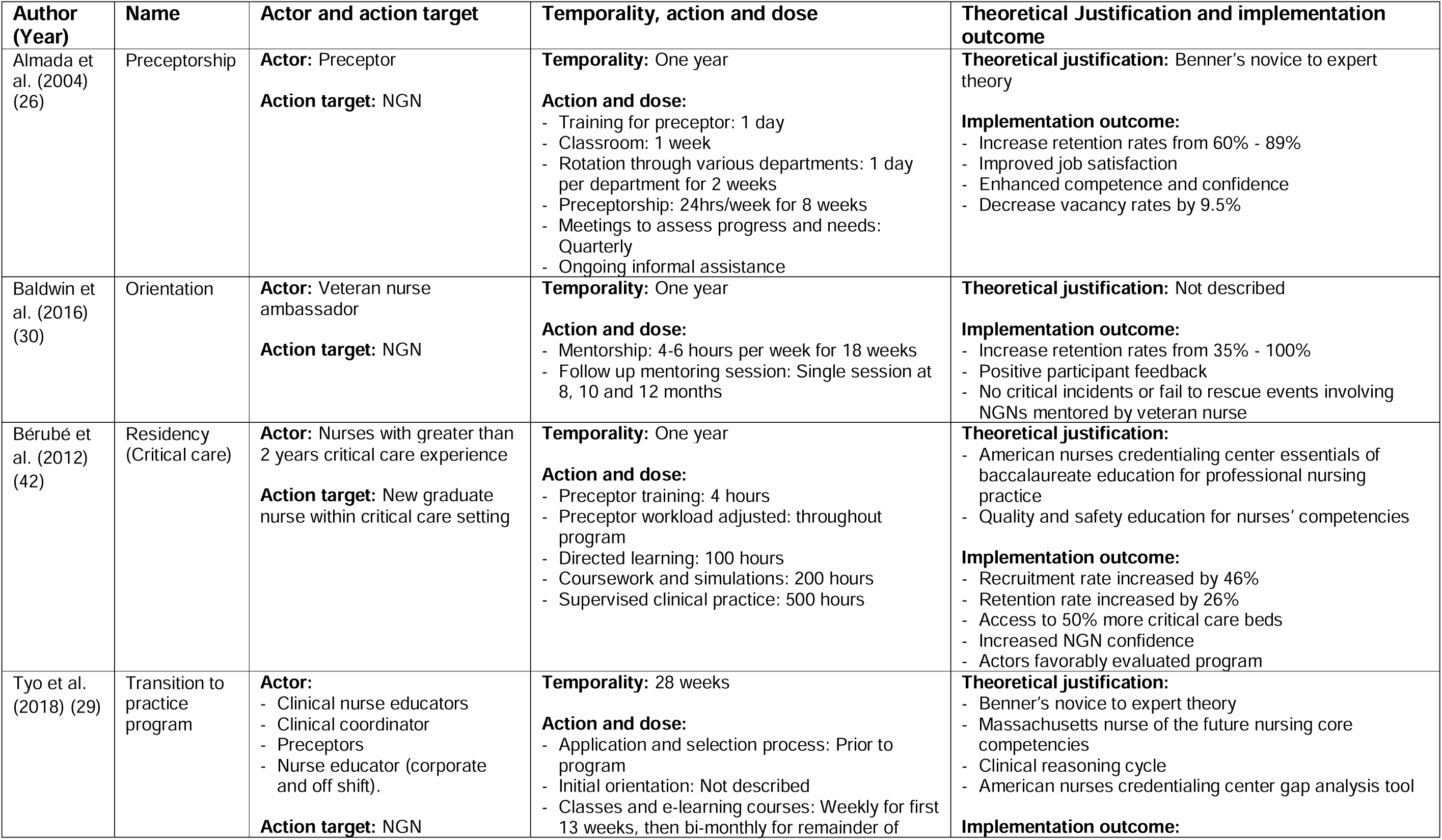
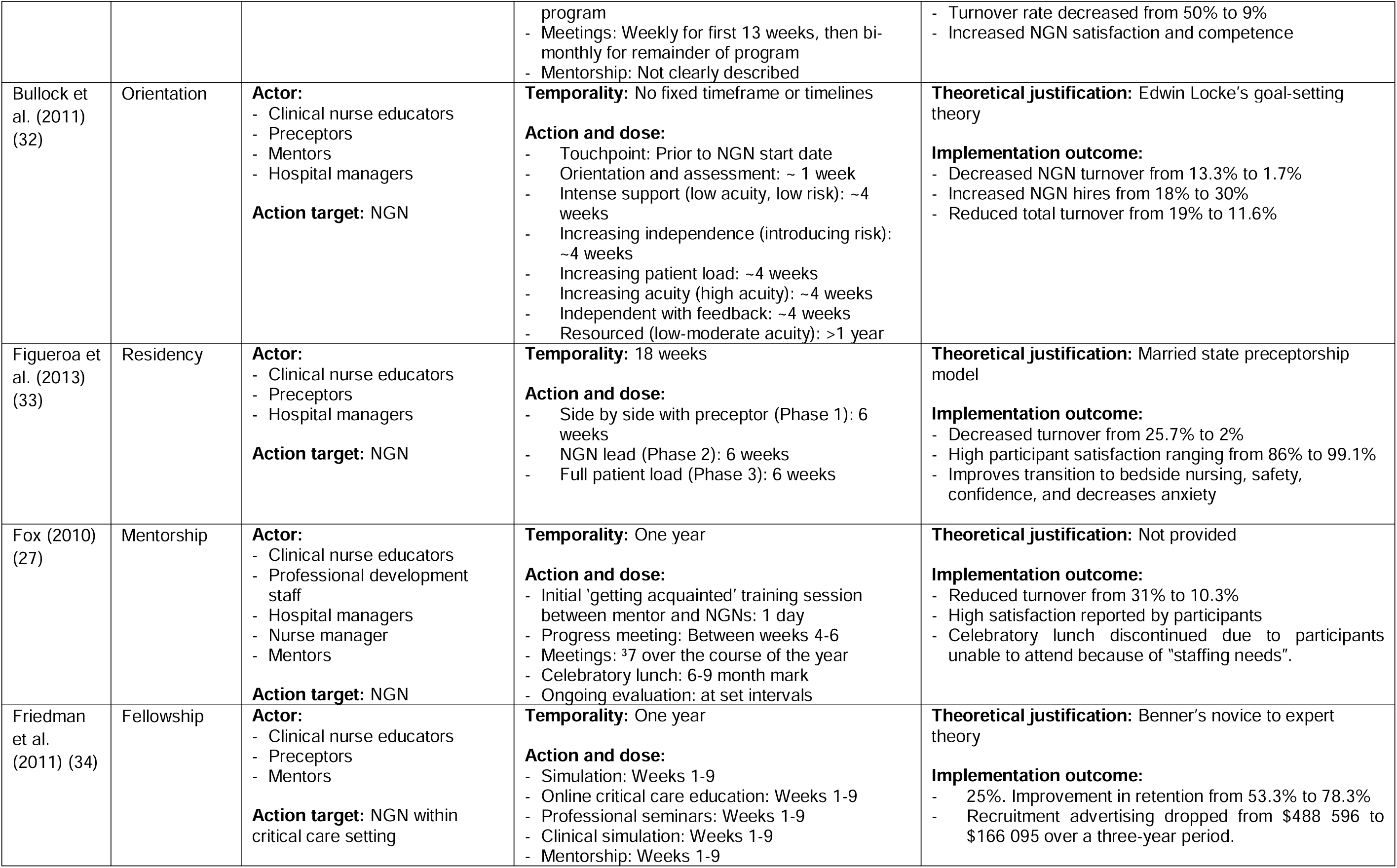
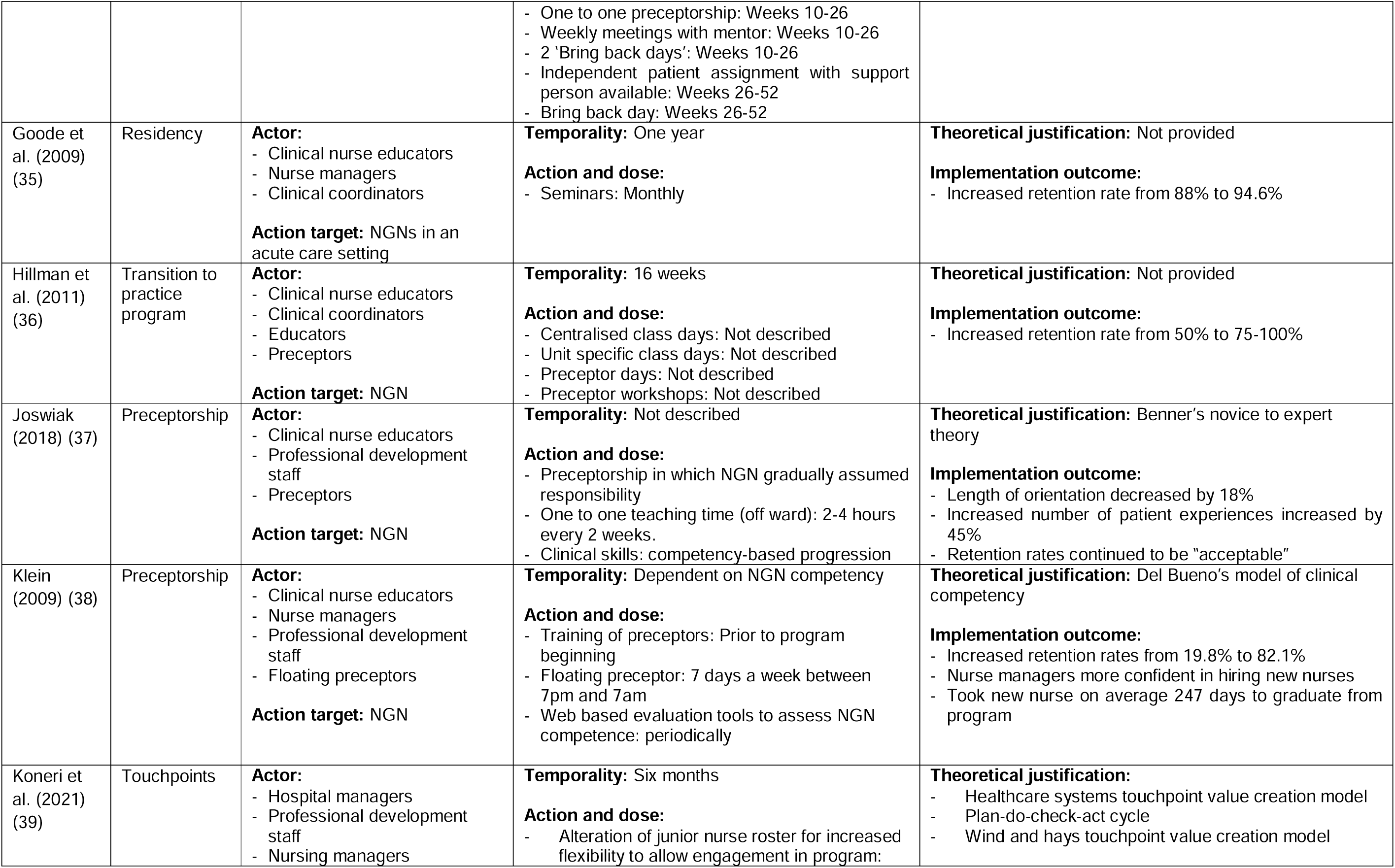
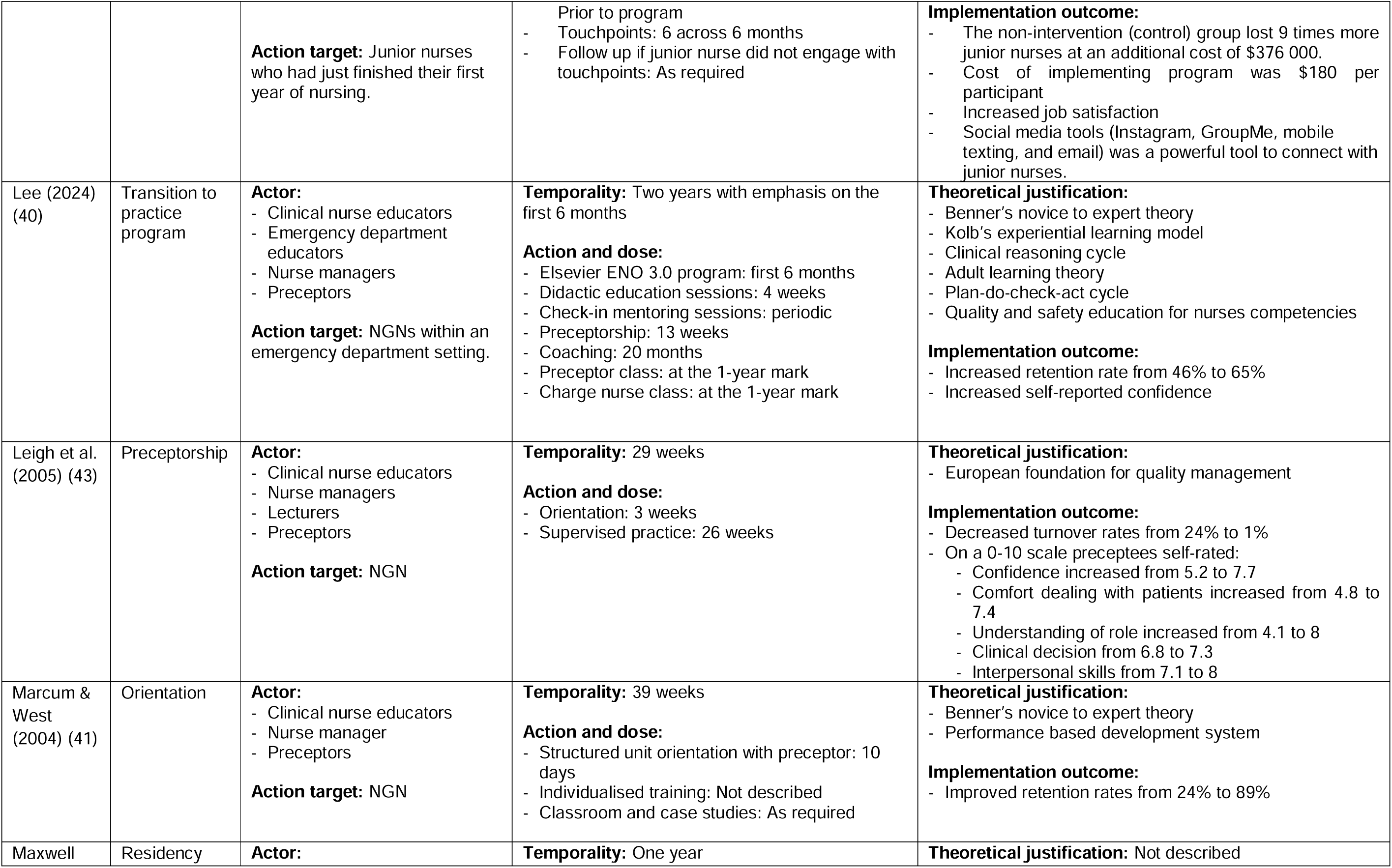
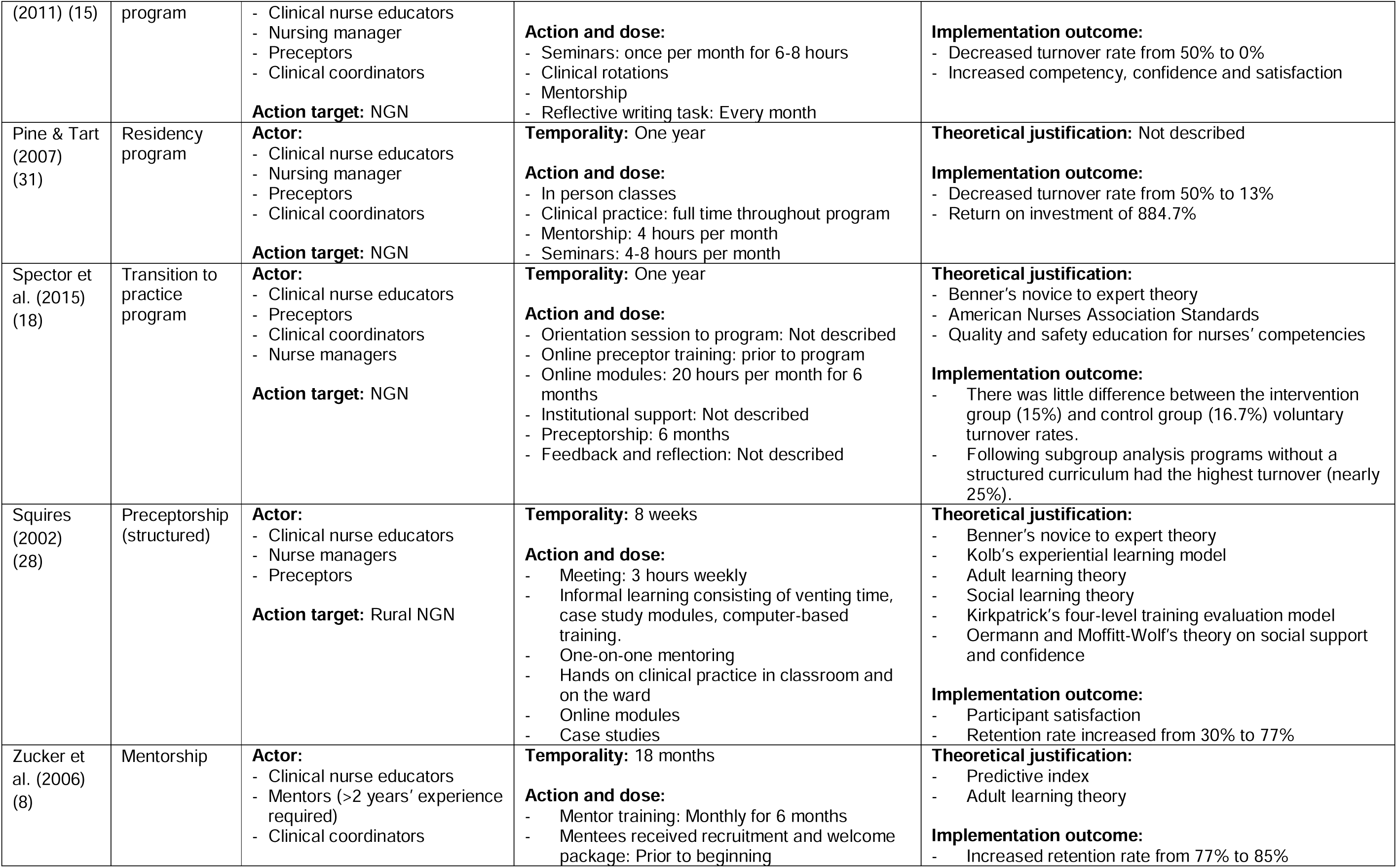
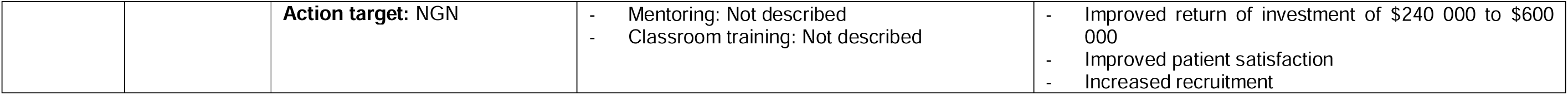
Characteristics of studies relating to the implementation science tool described by Proctor et al.

### Actors and the acted upon

Thirteen different job descriptors were utilised to describe individuals responsible for delivering interventions (Figure 3). Unsurprisingly, individuals responsible for engaging in face-to-face interactions, such as preceptors, clinical nurse educators, veteran nurse ambassadors, professional development and emergency department staff with the NGNs were the most commonly described ‘actors’. However, administrative and managerial staff (nurse managers, hospital managers, clinical coordinators) were also commonly involved in the delivery programs (Figure 3). Whilst many of the articles described interventions targeting NGNs from across their service, specialised interventions were identified within included studies to target NGNs within acute, critical care and emergency settings.

**Figure 3.**
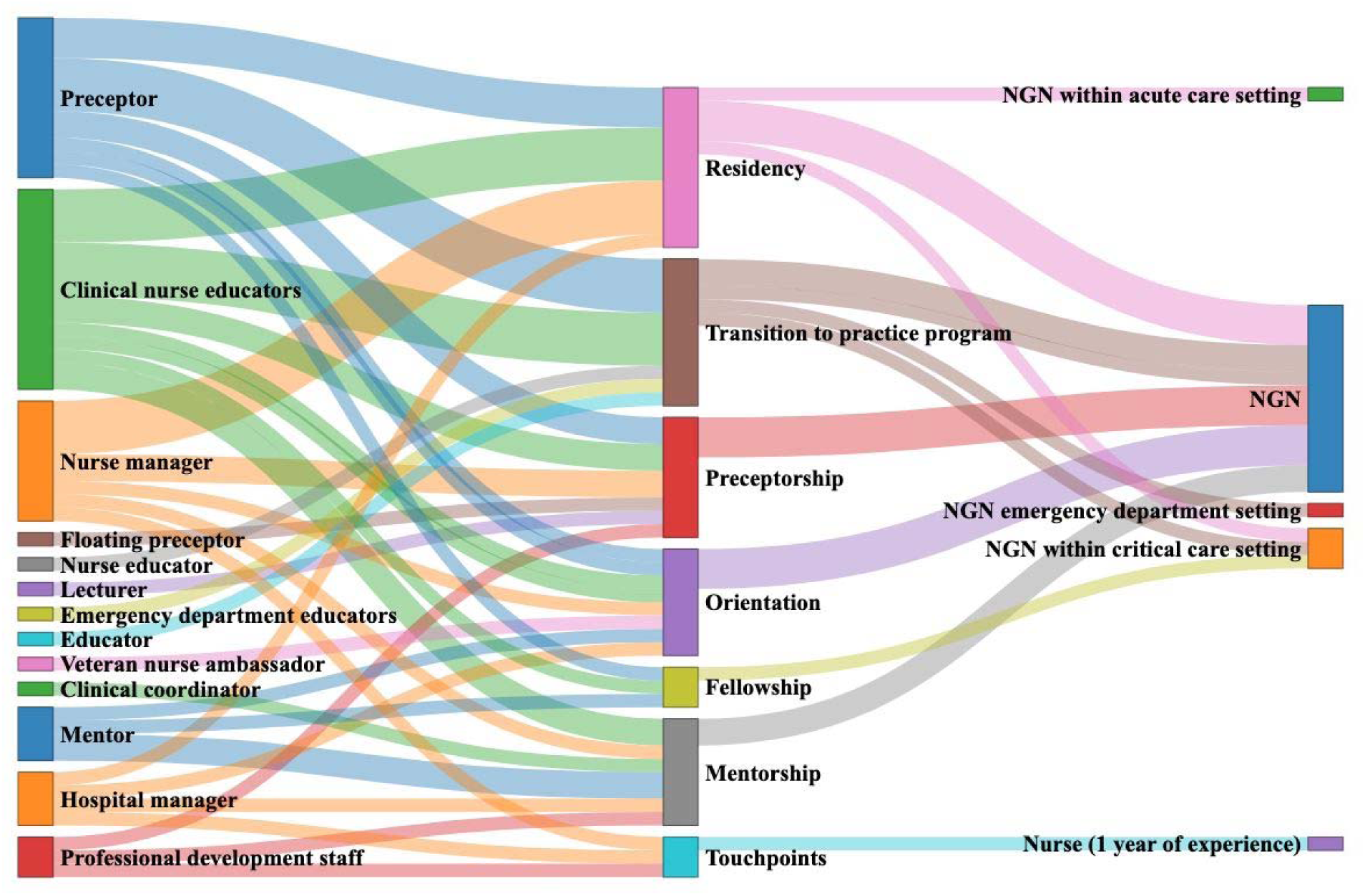
‘Actors’, their relationship to interventions and the ‘acted upon’

### Temporality

Intervention durations varied significantly across the programs reviewed, ranging from 8 weeks [(28)] to two years [(35)] (Figure 4.). Notably, two studies did not specify a fixed program length, instead tailoring the duration to individual participant competency levels [(15,38)]. Nine programs spanned a year and typically featured structured phases with ongoing support mechanisms [(15,18,26,27,30,31,34,35,42)]. One program focused on key interactions (touchpoints) within the first six months [(39)]. The shortest intervention was an 8-week preceptorship program [(28)], while more complex residency programs generally extended over a year [(28,30,36,37)], with one lasting 18 weeks [(32)]. Transition programs demonstrated the greatest variability, ranging from 16 weeks [(36)] to 28 weeks [(29)], to one year [(41)], with the longest extending to two years [(35)]. Preceptorship programs varied between 8 weeks [(28)] and one year [(26)], with two of these programs linking duration to participant competency [(15,38)]. Mentorship programs were less frequent and spanned between one year [(27) and 18 months [(8)].

**Figure 4.**
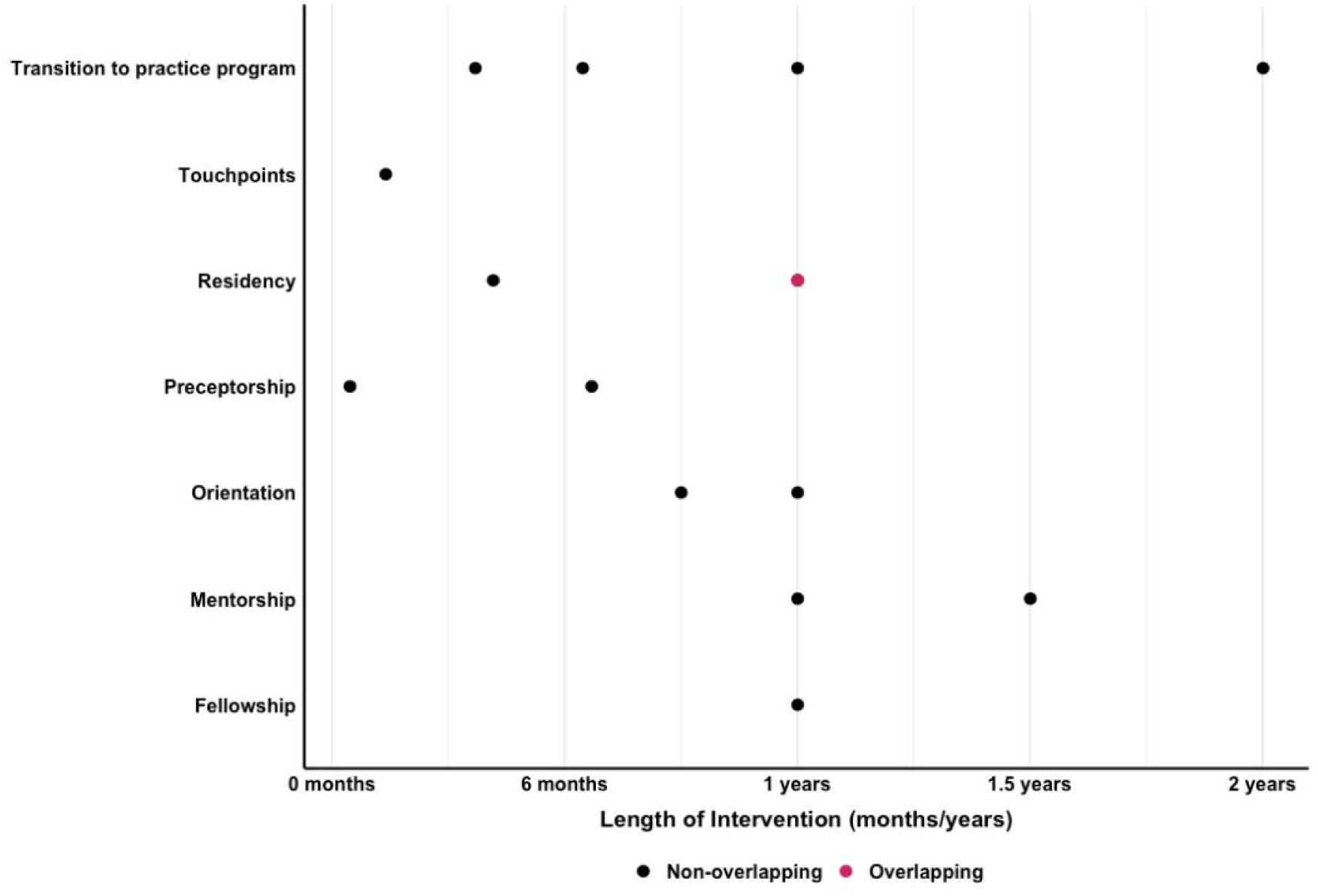
Length of interventions

### Action and dose

Interventions featured various delivery methods regarding what and for how long the actions were performed. All programs had an element of in person training or sessions, with eleven studies specifically describing mentoring and preceptorship [(18,28,30,32,34,36,37,39,40,43)]. Interestingly, these programs didn’t include training for their preceptors prior to implementation. Whereas some transition programs [(8,15,18,26–30,33,36–38,40–43)] included preceptor training prior to implementation. Furthermore, five studies within the programs awarded financial incentives to their preceptors during intervention implementation [(8,26,27,41,42)]. Other common themes included simulations [(29,30,34,38–40)] incorporating clinical practice [(15,29,30,32,35)] and focusing on reflective practice [(29,30,38)]. Institutional or professional support [(31,43)] and regular follow-up or feedback sessions [(18,28)] also featured regularly (Figure 5).

**Figure 5.**
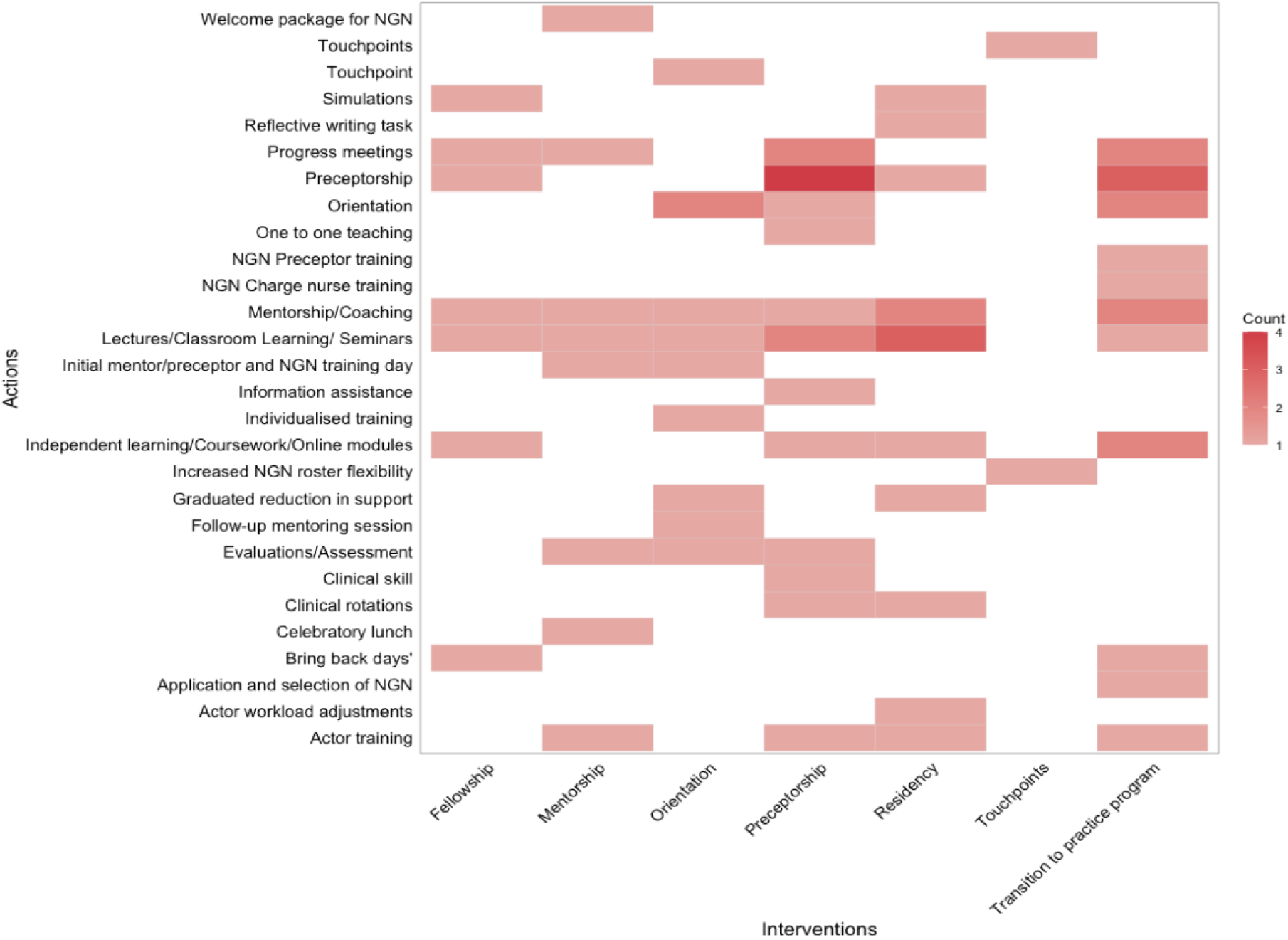
Actions associated with the interventions

### Theoretical Justification

There were twenty-six educational theories and models used within program designs. Benner’s Novice to Expert Theory [(18,26,28,34,37,40,41)] was used by four intervention types: orientation, fellowship, preceptorship and transition to practice programs. Programs that had a larger diversity in actions such as transition to practice programs and preceptorship also had a larger diversity in educational theories underpinning them. Other common appearances were Adult Learning Theory [(8,28,40)], Quality and Safety Education for Nurses competencies [(30,40,42,43)], Kolb’s Experiential Learning Model [(26,40)], Massachusetts Nurse of the Future Nursing Core Competencies [(29)], Clinical reasoning cycle [(29,40)], Casey Fink Graduate Nurse Experience Survey [(15,29)], and the Plan-Do-Check-Act Cycle [(39,40)]. Whilst others were used, they were less common (Figure 6).

**Figure 6.**
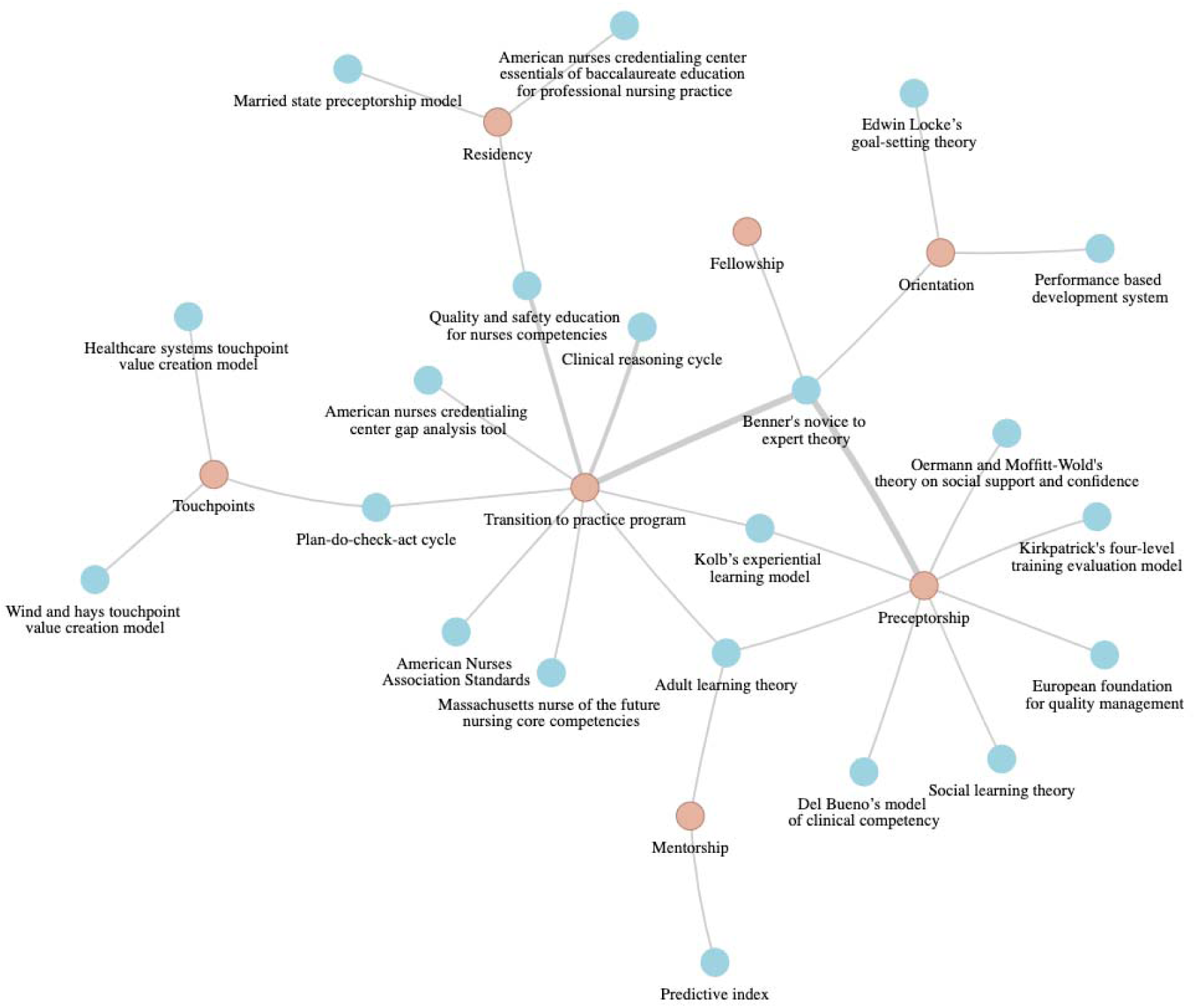
Theories informing interventions. Thickness of connection denotes the number of times utilised.

## Discussion

This scoping review sought to identify and analyse the key characteristics of interventions designed to retain early-career nurses (ECNs) within their first five years of practice, applying Proctor et al.’s (19) implementation science framework. The findings highlight the critical importance of structured mentorship programs, supportive leadership, and tailored retention strategies, particularly in light of the unique challenges posed by the COVID-19 pandemic.

### Structured mentorship as a foundation for retention

Mentorship programs emerged as a cornerstone for ECN retention, addressing challenges such as transition shock, inadequate preparedness, and lack of professional identity. Studies such as Vidal and Olley (44) and Young (45) highlight how structured mentorship enhances clinical performance, fosters a sense of belonging, and mitigates turnover intentions. Tailored mentorship approaches, such as pairing based on shared interests or specialties, offer additional benefits by enhancing engagement and communication (46).

The review identified the critical role of mentor preparation in supporting mentorship frameworks. Lindfors et al. (47) demonstrated that targeted educational interventions for preceptors significantly improved their ability to provide structured and consistent mentorship, resulting in enhanced professional competence among newly graduated nurses. This is further supported by Pohjamies et al. (48), who found that preceptor competence is directly associated with prior training and the ability to facilitate effective orientation. Competent preceptors are better equipped to provide tailored guidance, evaluate competencies, and foster an environment that supports skill acquisition and emotional resilience. Similarly, Lindfors et al. (49) highlighted the importance of structured orientation processes, where preceptor engagement significantly influences new graduate nurses’ perceptions of their clinical environment and role satisfaction. Collectively, these findings highlight that investing in preceptor training and ongoing support is essential for achieving both clinical and professional milestones, ultimately contributing to ECN retention.

Additionally, competency frameworks were identified as key components of effective mentorship programs. Lindfors et al. (47) demonstrated that interventions designed with clear competency goals, supported by structured preceptor education, significantly enhanced the professional competence of early-career nurses (ECNs). These findings align with the work of Rush et al. (16), who emphasised the importance of structured assessment and evaluation frameworks in formal transition-to-practice programs. However, the variability in how competencies are defined and evaluated across programs remains a challenge. For instance, some interventions implemented clear competency milestones, while others relied on informal observations (14). Developing standardised competency frameworks, alongside formal mentor training, could address these inconsistencies and improve program outcomes.

### The role of organisational culture and leadership

Supportive leadership and positive organisational culture were consistent themes across the literature. Leaders play a pivotal role in shaping work environments, addressing workload pressures, and providing emotional support. Studies like Djukic et al. (50) and Chargualaf et al. (51) emphasise the need for leadership-driven initiatives to create resilient and adaptive retention programs. The introduction of roles such as nurse retentionists, as suggested by Chargualaf et al. (51), offers a strategic approach to centralising recruitment, retention, and recognition efforts.

The findings also highlighted the importance of career development opportunities within retention interventions. Rush et al. (16) demonstrated that formal transition programs incorporating professional development and leadership training supported ECNs’ career readiness and long-term progression. By providing structured pathways to roles such as clinical leaders and shift managers, these initiatives contribute not only to short-term retention but also to workforce sustainability by building a pipeline for future leadership.

### Proctor’s framework and theoretical advances

Proctor et al.’s (19) framework provided a systematic lens for evaluating the components of retention interventions, including actors (who implements the intervention), actions (what is done), targets (who is affected), and dose (intensity and frequency). Findings from the scoping review revealed that the names of some interventions did not always reflect the actions undertaken, potentially leading to misinterpretation or underutilisation of effective strategies. Clearer and more descriptive nomenclature, as highlighted by Lindfors et al. (49), aligns with Proctor’s call for precise definitions of intervention components. Using this theoretical approach ensures that interventions are both replicable and accessible, enabling a more nuanced evaluation of their impact on retention outcomes.

The scoping review identified challenges related to intervention length and structure. Flexible programs offer adaptability to individual learning needs but may lead to inconsistencies in skill acquisition. In contrast, fixed timelines ensure uniform outcomes but may not account for individual variations in learning speed. The balance between flexibility and standardisation, as highlighted by Chappell and Richards (52), is crucial for tailoring interventions to meet diverse ECN needs while maintaining consistent program outcomes.

### Emerging Issues and Future Directions

#### 1. Specialised Interventions for Critical Care Settings

The scoping review highlighted the growing demand for specialised interventions tailored to high-stress environments, such as critical care. These settings pose unique challenges for ECNs, including high patient acuity and emotional strain, which can exacerbate burnout for both ECNs and senior nurses. Interventions must account for these dynamics to prevent attrition and avoid overburdening experienced staff.

#### 2. Lessons from the COVID-19 Pandemic

The pandemic exposed significant vulnerabilities in traditional retention strategies, such as the disruption of clinical placements and the shift to remote learning. This led to a lack of preparedness and confidence among ECNs, as noted in studies like Chargualaf et al. (2023). Enhanced transition-to-practice (TTP) programs that integrate simulation-based learning and mental health resources are essential for addressing these gaps.

#### 3. Addressing Research Gaps

This review identifies critical gaps in the literature, including the need for longitudinal studies that assess the scalability and adaptability of retention interventions across diverse settings. Few studies explore retention strategies for ECNs in rural, remote, or non-acute care environments. Additionally, limited research examines the direct impact of mentorship on organisational metrics, such as patient outcomes and financial sustainability. Addressing these gaps is vital for refining evidence-based practices and ensuring the broader applicability of retention strategies.

## Conclusion

This review highlights the critical role of structured mentorship, supportive leadership, and tailored retention interventions in addressing the challenges faced by ECNs. By leveraging Proctor’s framework and lessons from the COVID-19 pandemic, healthcare organisations can design and implement adaptive retention strategies that foster a resilient nursing workforce. Addressing identified gaps and prioritising the mental health and professional development of ECNs will be essential for ensuring workforce sustainability and improving patient care outcomes.

## Data Availability

All data produced in the present study are available upon reasonable request to the authors

## Acknowledgements

The authors would like to thank Bond University for providing the resources necessary for conducting this research. This review is part of the coursework contributing towards the Award of Master of Healthcare Innovations Degree for Krishangi White.

## Author’s contributions

CRediT author statement – **Krishangi White**: Conceptualisation, Data curation, Formal analysis, Investigation, Methodology, Project administration, Writing – original draft. **Adrian Goldsworthy**: Conceptualisation, Data curation, Formal analysis, Investigation, Methodology, Project administration, Software, Resources, Supervision, Validation, Visualisation, Writing – review & editing. **Sarah Bateup**: Investigation, Methodology. **Neil Meigh**: Conceptualisation, Investigation, Methodology, Project administration, Resources, Supervision, Writing – review & editing.

## Availability of data and materials

All data produced in the present study are available upon reasonable request to the authors.

## Declarations

### Ethics approval and consent to participate

Not applicable.

### Consent for publication

Not applicable.

## Appendix 1

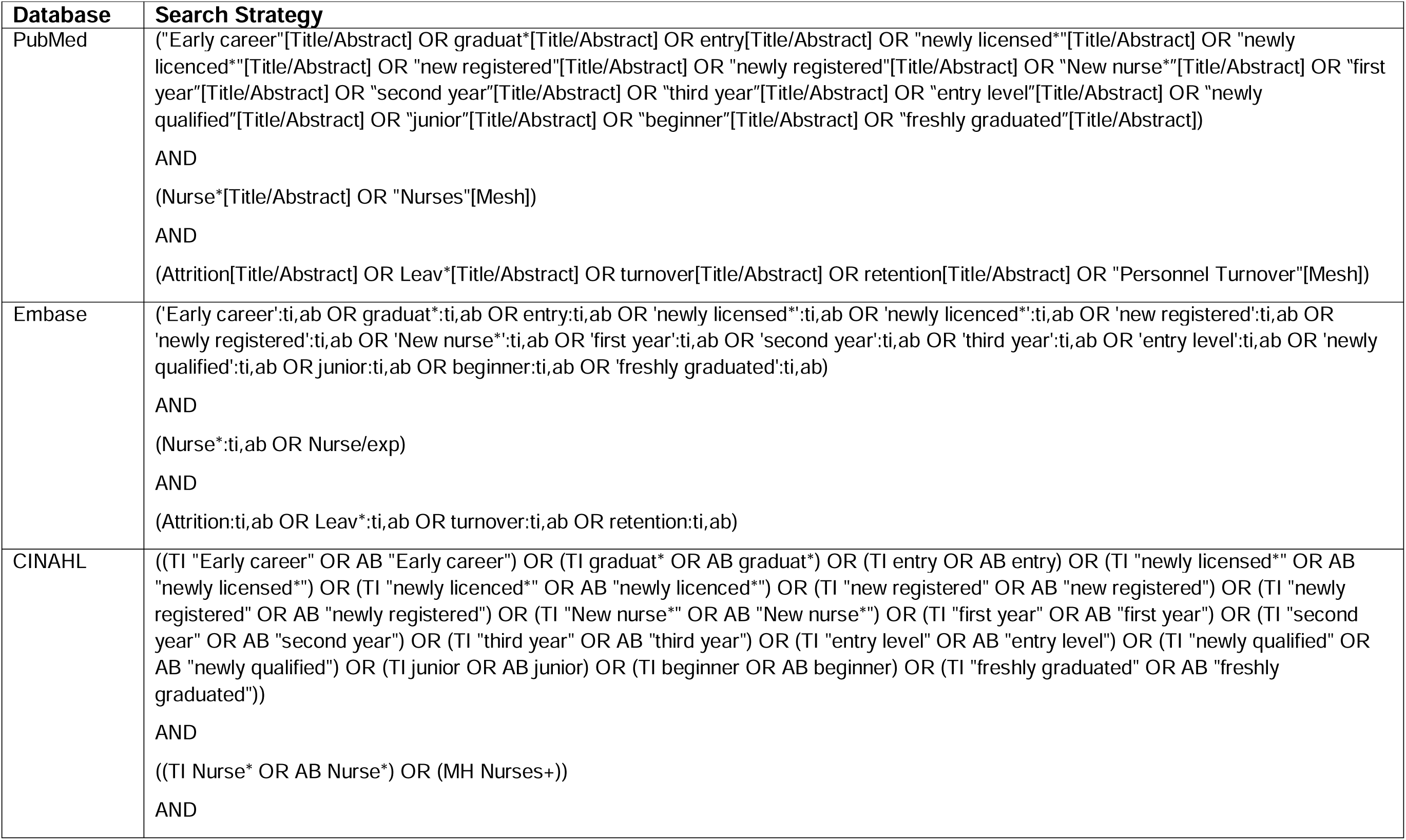
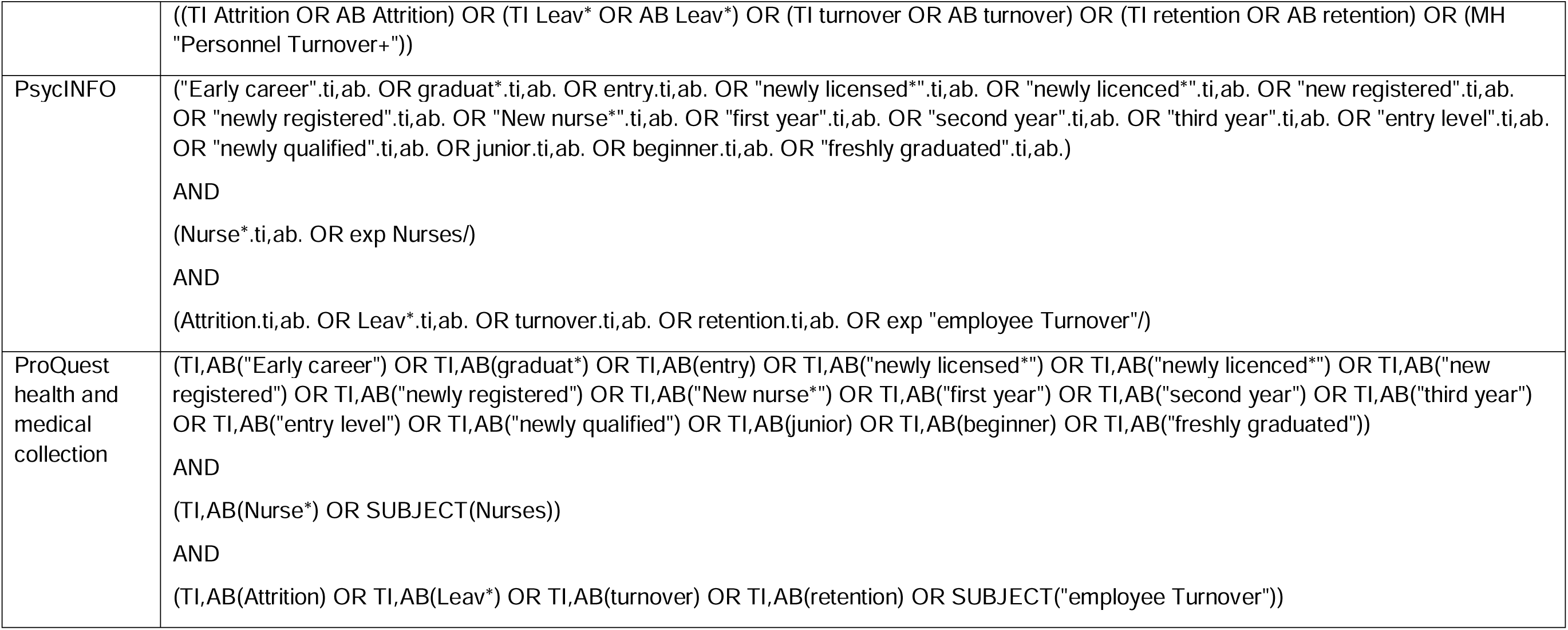

## References

1. Brewer CS, Kovner CT, Greene W, Tukov-Shuser M, Djukic M. Predictors of actual turnover in a national sample of newly licensed registered nurses employed in hospitals. J Adv Nurs. 2012;68(3):521–38.

2. Kovner CT, Brewer CS, Fatehi F, Jun J. What does nurse turnover rate mean and what is the rate? Policy Polit Nurs Pract. 2014;15(3–4):64–71.

3. Australian Institute of Health and Welfare [Internet]. 2024 [cited 2024 Dec 17]. Workforce Overview. Available from: https://www.aihw.gov.au/reports-data/health-welfare-services/workforce/overview

4. Nearly half of Queensland nurses and midwives considering leaving within 12 months - ANMJ [Internet]. 2024 [cited 2024 Dec 17]. Available from: https://anmj.org.au/nearly-half-of-queensland-nurses-and-midwives-considering-leaving-within-12-months/

5. Brook J, Aitken L, Webb R, MacLaren J, Salmon D. Characteristics of successful interventions to reduce turnover and increase retention of early career nurses: A systematic review. Int J Nurs Stud. 2019 Mar;91:47–59.

6. Vázquez-Calatayud M, Eseverri-Azcoiti MC. Retention of newly graduated registered nurses in the hospital setting: A systematic review. J Clin Nurs. 2023;32(19–20):6849–62.

7. Chen F, Liu Y, Wang X, Dong H. Transition shock, preceptor support and nursing competency among newly graduated registered nurses: A cross-sectional study. Nurse Educ Today. 2021 Jul;102:104891.

8. Zucker B, Goss C, Williams D, Bloodworth L, Lynn M, Denker A, et al. Nursing retention in the era of a nursing shortage: Norton Navigators. J Nurses Staff Dev. 2006;22(6):302–6.

9. Chan GK, Bitton JR, Allgeyer RL, Elliott D, Hudson LR, Burwell PM. The impact of COVID-19 on the nursing workforce: a national overview. Online J Issues Nurs. 2021;26(2):1–17.

10. Casey K, Fink RR, Krugman AM, Propst FJ. The Graduate Nurse Experience: JONA J Nurs Adm. 2004 Jun;34(6):303–11.

11. Fink R, Krugman M, Casey K, Goode C. The Graduate Nurse Experience: Qualitative Residency Program Outcomes. JONA J Nurs Adm. 2008 Jul;38(7/8):341–8.

12. Weller-Newton JM, Murray M, Phillips C, Laging B, McGillion A. Transition to Practice Programs in Nursing: A Rapid Review. J Contin Educ Nurs. 2022 Oct;53(10):442–50.

13. Brown JA, Capper T, Hegney D, Donovan H, Williamson M, Calleja P, et al. Individual and environmental factors that influence longevity of newcomers to nursing and midwifery: a scoping review. JBI Evid Synth. 2024;22(5):753–89.

14. Rush KL, Janke R, Duchscher JE, Phillips R, Kaur S. Best practices of formal new graduate transition programs: An integrative review. Int J Nurs Stud. 2019 Jun;94:139–58.

15. Maxwell KL. The implementation of the UHC/AACN New Graduate Nurse Residency Program in a community hospital. Nurs Clin North Am. 2011;46(1):27–33.

16. Rush KL, Adamack M, Gordon J, Lilly M, Janke R. Best practices of formal new graduate nurse transition programs: An integrative review. Int J Nurs Stud. 2013 Mar;50(3):345–56.

17. Masso M, Sim J, Halcomb E, Thompson C. Practice readiness of new graduate nurses and factors influencing practice readiness: A scoping review of reviews. Int J Nurs Stud. 2022 May;129:104208.

18. Spector N, Blegen MA, Silvestre J, Barnsteiner J, Lynn MR, Ulrich B, et al. Transition to Practice Study in Hospital Settings. J Nurs Regul. 2015;5(4):24–38.

19. Proctor EK, Powell BJ, McMillen JC. Implementation strategies: recommendations for specifying and reporting. Implement Sci. 2013;8(1):1–11.

20. Peters MD, Marnie C, Tricco AC, Pollock D, Munn Z, Alexander L, et al. Updated methodological guidance for the conduct of scoping reviews. JBI Evid Synth. 2020;18(10):2119–26.

21. Tricco AC, Lillie E, Zarin W, O’Brien KK, Colquhoun H, Levac D, et al. PRISMA extension for scoping reviews (PRISMA-ScR): checklist and explanation. Ann Intern Med. 2018;169(7):467–73.

22. Clark JM, Sanders S, Carter M, Honeyman D, Cleo G, Auld Y, et al. Improving the translation of search strategies using the Polyglot Search Translator: a randomized controlled trial. J Med Libr Assoc JMLA. 2020;108(2):195.

23. Gotschall T. EndNote 20 desktop version. J Med Libr Assoc JMLA. 2021;109(3):520.

24. Kellermeyer L, Harnke B, Knight S. Covidence and rayyan. J Med Libr Assoc JMLA. 2018;106(4):580.

25. Microsoft Excel [Internet]. (Publisher): Microsoft Corporation; 2024. Available from: https://www.microsoft.com/microsoft-365

26. Almada P, Carafoli K, Flattery JB, French DA, McNamara M. Improving the retention rate of newly graduated nurses. J Nurses Staff Dev. 2004;20(6):268–73.

27. Fox KC. Mentor program boosts new nurses’ satisfaction and lowers turnover rate. J Contin Educ Nurs. 2010;41(7):311–6.

28. Squires A. New graduate orientation in the rural community hospital. J Contin Educ Nurs. 2002;33(5):203–9.

29. Tyo MB, Gundlach M, Brennan C, Esdale L, Knight A, Provencher S, et al. Leading the charge: Achievement of national accreditation for a nurse residency program. J Nurses Prof Dev. 2018;34(5):270–6.

30. Baldwin KM, Black DL, Normand LK, Bonds P, Townley M. Integrating Retired Registered Nurses Into a New Graduate Orientation Program. Clin Nurse Spec J Adv Nurs Pract. 2016;30(5):277–83.

31. Pine R, Tart K. Return on investment: benefits and challenges of baccalaureate nurse residency program. Nurs Econ. 2007;25(1):13–8, 39, 3.

32. Bullock LM, Groff Paris L, Terhaar M. Designing an outcome-focused model for orienting new graduate nurses. J Nurses Staff Dev. 2011;27(6):252–8.

33. Figueroa S, Bulos M, Forges E, Judkins-Cohn T. Stabilizing and retaining a quality nursing work force through the use of the married state preceptorship model. J Contin Educ Nurs. 2013;44(8):365–73.

34. Friedman MI, Cooper AH, Click E, Fitzpatrick JJ. Specialized new graduate rn critical care orientation: retention and financial impact. Nurs Econ. 2011;29(1):7–14.

35. Goode CJ, Lynn MR, Krsek C, Bednash GD. Nurse residency programs: An essential requirement for nursing. Nurs Econ. 2009;27(3):142.

36. Hillman L, Foster RR. The impact of a nursing transitions programme on retention and cost savings. J Nurs Manag. 2011;19(1):50–6.

37. Joswiak ME. Transforming Orientation Through a Tiered Skills Acquisition Model. J Nurses Prof Dev. 2018;34(3):118–22.

38. Klein GS. Beyond orientation: Developing and retaining new graduate nurses. Nurs Manag (Harrow). 2009;40(1):10–3.

39. Koneri L, Green A, Gilder RE. Touchpoints: A Business Strategy to Retain New Graduate Nurses. J Nurs Adm. 2021;51(7–8):401–8.

40. Lee MMD. Improving New Graduate Nurse Retention With a Transition to Emergency Nursing Practice Program. J Emerg Nurs. 2024;50(2):178–86.

41. Marcum EH, West RD. Structured orientation for new graduates: A retention strategy. J Nurses Staff Dev. 2004;20(3):118–24.

42. Bérubé M, Valiquette MP, Laplante E, Lepage I, Belmonte A, Tanguay N, et al. Nursing residency program: a solution to introduce new grads into critical care more safely while improving accessibility to services. Nurs Leadersh Tor Ont. 2012;25(1):50–67.

43. Leigh JA, Douglas CH, Lee K, Douglas MR. A case study of a preceptorship programme in an acute NHS Trust -- using the European Foundation for Quality Management tool to support clinical practice development. J Nurs Manag. 2005;13(6):508–18.

44. Vidal JAM, Olley R. Systematic Literature Review of The Effects Of Clinical Mentoring On New Graduate Registered Nurses’ Clinical Performance, Job Satisfaction And Job Retention. Asia Pac J Health Manag. 2021 Dec 13;16(4):70–82.

45. Young J. Reach Them to Keep Them: The Effects of Mentorship on New - Google Search [Internet]. Georgia State University; 2024 [cited 2024 Nov 29]. Available from: 10.57709/36947559

46. Mínguez Moreno I, González de la Cuesta D, Barrado Narvión MJ, Arnaldos Esteban M, González Cantalejo M. Nurse Mentoring: A Scoping Review. Healthc Basel Switz. 2023 Aug 15;11(16):2302.

47. Lindfors K, Flinkman M, Kaunonen M, Huhtala H, Paavilainen E. New graduate registered nurses’ professional competence and the impact of preceptors’ education intervention: a quasi-experimental longitudinal intervention study. BMC Nurs. 2022 Dec 16;21(1):360.

48. Pohjamies N, Haapa T, Kääriäinen M, Mikkonen K. Nurse preceptors’ orientation competence and associated factors—A cross-sectional study. J Adv Nurs. 2022 Dec;78(12):4123–34.

49. Lindfors K, Kaunonen M, Huhtala H, Paavilainen E. Newly graduated nurses’ evaluation of the received orientation and their perceptions of the clinical environment: An intervention study. Scand J Caring Sci. 2022 Mar;36(1):59–70.

50. Djukic M, Padhye N, Ke Z, Yu E, McVey C, Manuel W, et al. Associations Between the COVID-19 Pandemic and New Nurses’ Transition to Practice Outcomes: A Multi-site, Longitudinal Study. J Nurs Regul. 2023 Apr;14(1):42–9.

51. Chargualaf KA, Bourgault A, Torkildson C, Graham-Clark C, Nunez S, Barile LT, et al. Retaining new graduate nurses: Lessons learned from the COVID-19 pandemic. Nurs Manag (Harrow). 2023 Sep 1;54(9):26–34.

52. Chappell KB, Richards KC. New graduate nurses, new graduate nurse transition programs, and clinical leadership skill: a systematic review. J Nurses Prof Dev. 2015;31(3):128–37.

